# Improving Mortality Surveillance through Notification of Death at Mbale Regional Referral Hospital, Uganda, October 2023–April 2024

**DOI:** 10.1101/2024.12.09.24318739

**Authors:** Innocent Ssemanda, Edith Namulondo, Carol Naziri, Stephen Obbo, Benon Kwesiga, Richard Migisha, Lilian Bulage, Job Morukileng, Alex Riolexus Ario

## Abstract

**Background:** Accurate mortality reporting is crucial for monitoring population health, detecting disease outbreaks, and informing health policies. However, the implementation of medical certification of cause of death remains low in Uganda, with only 3.2% of health facility deaths being notified to the Ministry of Health. Using a quality improvement approach, we aimed to improve mortality reporting through medical certification of cause of death at Mbale Regional Referral Hospital (MRRH) in Uganda from 1% to 80% within 6 months.

**Methods:** We purposively selected MRRH as one of five regional referral hospitals with the lowest death notifications (0%-20%) during 2022 and 2023. We adopted the existing quality improvement team, which includes medical and non-medical personnel. Focus group discussions identified challenges that informed the root cause analysis. Using the Plan-Do-Study-Act (PDSA) cycle, we generated change ideas (interventions) to address these bottlenecks. We monitored the progress of the interventions with process indicators (number of mentorship sessions conducted, number of review meetings held) and an outcome indicator (proportion of deaths occurring in the hospital notified through the District Health Information System version 2 (DHIS2)) for 6 months. We tracked notifications monthly and analyzed the trend at six months using the Mann-Kendall test.

**Results:** We conducted 4/6 (67%) mentorship sessions and 7/19 (38%) review meetings and trained 32/50 (64%) nurses. The qualitative findings highlighted key challenges, including lack of knowledge and training, competing priorities and workload, resource constraints, undervaluing the importance of mortality reporting, failure to follow guidelines, and heavy workloads. The interventions included training and mentorship sessions for the staff on properly completing the death notification form, adopting a standardized process for form completion, and conducting bi-monthly review meetings. The proportion of deaths notified through DHIS2 from November 2023 to April 2024 increased from 17% to 65% (p=0.01).

**Conclusion:** Training of staff, adoption of a standard protocol on notification, and routine review meetings could facilitate death notification and improve mortality surveillance in Uganda enabling more accurate resource allocation for mortality prevention. The target was not met probably because all the staff were not trained, and the review meetings were sub-optimal.

## 1.0 Background

Medical notification and certification of Cause of Death (MCCoD) involves authorized medical personnel determining and documenting the cause of death to maintain accurate mortality records. These records are essential for monitoring health indices, detecting disease outbreaks, assessing health interventions, and strengthening the civil registration and vital statistics (CRVS) system for informed health policy and planning [1, 2].

Death registration varies from close to 100% in the developed countries of Europe and America to around 50% in the Asia-Pacific region and less than 10% among African countries including Uganda [3]. In 2022, the World Bank estimated the crude mortality rate in Uganda to be at 6.4 per 1000 people [4]. This means Uganda registered more than 283,000 deaths in that year. About 159,996 (67%) of these deaths occurred in the communities and the other 93,654 (33%) occurred at the health facility. According to the District Health Information System 2 (DHIS2), only 63,499 (69%) of the health facility deaths were captured and only 2,060 (3.2%) of these were notified to the Ministry of Health (MoH) with a valid MCCoD underlying cause.

With the use of MCCoD data, the World Health Organization (WHO), formulated guidelines for the International Classification of Disease (ICD), designed to promote international comparability in the collection, processing, classification, and presentation of morbidity and mortality data. These guidelines include a form for medical certification of cause of death which form\requires certifying physicians to record a pathophysiological sequence of clinical conditions leading to death, their durations, and other contributory causes [2, 5].

Of the deaths recorded in DHIS2, 34% (21,831) occurred in 21 public facilities, including 4 national referral hospitals and 17 regional referral hospitals. However, only 369 of these cases (1.2%) had a valid Medical Certification of Cause of Death (MCCoD). This indicates that most deaths in these health facilities are not properly documented or registered, leading to a significant loss of vital information about mortality causes and patterns in the region. Such gaps also undermine the quality of healthcare services and impact the effective allocation of resources.

A continuous quality improvement (CQI) approach can be an effective strategy for improving death notification compliance. Continuous Quality improvement is a progressive process of measurable improvement in the efficiency and effectiveness of performance outcomes in services or processes, to improve patient care. The CQI approach has been successfully used to improve performance practices in various settings. By employing a plan-do-study-act (PDSA) cycle, low-cost, high-impact interventions with continuous monitoring of mortality reporting can be applied iteratively in a clinical setting to improve mortality reporting. This iterative process allows for the development and testing of solutions, followed by evaluation and refinement, ultimately leading to sustainable improvements in mortality reporting [6, 7].

In Uganda, the Medical Notification and Certification of Cause of Death (MCCoD) was rolled out by the World Health Organization (WHO) in 2021. Two years later, in 2023, the overall rate of death certification remains low at 11%. This underscores the need for improvement, which this project undertook through a continuous quality improvement approach. Mbale Regional Referral Hospital (MRRH) is the largest and the busiest hospital in the Bugisu subregion, Eastern Uganda, reporting up to 259 deaths in DHIS2, yet its notification remains below 5%. This quality improvement project aimed to identify and analyze the factors associated with low death notification rates, address these factors, and highlight the successful implementation of improved death notification practices at MRRH.

## 2.0 Methods

### 2.1 Quality improvement setting and design

We conducted the project at MRRH, a government-owned and funded hospital. It is located 250km east of Kampala in the centre of Mbale City. The hospital serves over 4.6 million people in 16 surrounding districts. Started in 1924 as a health Centre, it has since grown to a 450-bed capacity regional referral hospital [8]. The hospital has ample capacity to document death notifications since it requires only one nurse on duty to fill out the form following a patient’s death.

We employed continuous quality improvement using a Plan-Do-Study-Act (PDSA) cycle where several challenges to death notification were identified and ranked, and solutions were proposed during the baseline assessment. The project was carried out in three phases i.e. baseline, intervention implementation for six months, and evaluation at the end.

Analysis of DHIS2 data a year before the project inception (September 2022 to August 2023), revealed that only 1% of the regional referral recorded deaths were being notified of about 200 deaths every month on average.

### 2.2 Pre-intervention and inception

We held an inception meeting with the Hospital Director and the heads of department of MRRH to introduce the project focus and identify resourceful persons.

### 2.3 Formation and training of the Quality Improvement Team

We held a meeting with the clinical and nursing staff and adopted an existing continuous quality improvement (CQI) team, adding a few members to coordinate the project activities. The existing CQI team was originally selected based on qualities such as enthusiasm, interest in improving quality, influence among co-workers, and ability to get things done.

We trained the team on the quality improvement processes and mortality surveillance to help them identify and prioritize problems, develop a clear aim statement, identify what to measure and change ideas and implement a series of PDSA cycles to develop and test the changes.

### 2.4 Baseline assessment

A baseline review was conducted to determine the status of the certified deaths in MRRH. Available data from the inpatient deaths reported and the notified death was extracted and analyzed. Self-administered questionnaires were also conducted to identify gaps that needed to be addressed. This information helped the CQI team to identify and prioritize key improvement areas.

### 2.5 Problem identification

The qualitative component of the study involved a focused group discsuion with key stakeholders to understand the factors contributing to the low mortality reporting rates at MRRH. The interviews explored the perspectives and experiences of hospital staff in completing the HMIS 100 death notification form and the challenges they faced

#### 2.5.1 Problem prioritization

The CQI team used a prioritization matrix for the three problems identified. Based on their experience, the team assigned points from 1-5 to each of the following prioritization aspects.

a. *Importance to the deceased’s next of kin*: A score of 1 means the problem is not important, and a score of 5 means it is vitally important.
b. *Affordable in terms of time and resources:* A score of 1 means fixing the problem will take a lot of resources and time, and a score of 5 means it is very affordable and takes a reasonable time to fix.
c. *Easy to measure*: A score of 1 means the problem is not measurable, and a score of 5 means it is very easy to measure using existing tools.
d. *Under the control of the team:* A score of 1 means the problem is not under the team’s control, and a score of 5 means it is entirely under the team’s control.

### 2.6 Setting the QI aims

Based on the prioritization matrix, the team proposed to fill the HMIS form 100 notification sections as soon as the death occurs. From the problem, we found an aim statement, specifically stating what we wanted to improve, who would benefit (the specific patient population), how much improvement we wanted, and by when (the timeline).

### 2.7 Defining and describing the problem

The mortality/death notification form (HMIS form 100) has 2 main sections. Section 1 has two parts (1a and 1b), both capturing the demographics of the deceased.

Section 2 is divided into frames. Frame A, having parts 1 and 2, captures medical data, and Frame B captures other medical data. Lastly, the form has a declaration section, where the medical practitioner certifying the death signs.

To notify, once death has occurred, the nurse must immediately complete Part 1a, part 1b, and Frame B of the HMIS 100. The Medical Records Officer (MRO) should get the deceased’s file containing the HMIS 100 with the filled-in part 1a, part 1b, and Frame B. The MRO enters the information on Part 1a, part 1b, and Frame B into DHIS2. Once the information is entered into DHIS2, the MRO returns the filled HMIS form 100 to the ward for the Medical Doctor to certify the death, fill the sequence of cause of death in Frame A, and complete the other sections of the form.

### 2.8 Aim statement

The QI project aimed to increase the proportion of deaths occurring in the hospital that are notified through DHIS2 from 17% to 80% starting October 2023 through April 2024.

### 2.9 Root cause analysis

We used a fishbone analysis technique to identify the reasons why the death notification form 100 is not filled immediately after death. The root causes were categorized as personnel, capacity, environment, monitoring, and guidelines/reminders.

### 2.10 Intervention

This phase involved developing and testing a set of interventions to address the factors identified in the baseline assessment. The interventions included four-day training sessions for the CQI team conducted by the principal investigator, two quarterly supervisory visits, and two Continuous Medical Education (CME) sessions. The teams were trained in quality improvement principles and processes, during which they identified factors affecting mortality certification and coding, selected those they could address, developed clear action plans, identified the quality improvement tools to use, and implemented a series of PDSA cycles to develop and test the solutions. The interventions were implemented simultaneously from the beginning of the project, and the PDSA framework was used to make minor adjustments as needed along the way.

The key aspects of this approach were measuring and monitoring performance outcomes, identifying and addressing gaps or inefficiencies, implementing targeted, evidence-based interventions, continuously evaluating and refining the interventions through the PDSA cycle, and maintaining a focus on improving patient care and outcomes.

Based on the root causes, the QI team used their experience to suggest related changes to address the root causes and evaluated them using the PDSA cycle. The proposed changes maintained the acceptable standards of service delivery as per MoH guidelines. The team developed an implementation plan for the proposed changes shown in Table 1.

**Table 1.** Implementation plan (Plan-do-study-act) for the suggested changes to address the root causes for failure to fill form 100 immediately after death at Mbale Regional Referral Hospital, 2023-2024.

|  |  |  |
| --- | --- | --- |
| **Plan** | The proposed changes? | 1 Conducted a mentorship session for all nursing staff on the importance of filling out HMIS form 100 immediately after death<br>2 Developed a standardized process for filling HMIS form 100, including clear roles and responsibilities for each staff member<br>3 Ensured that all necessary supplies (e.g., MHIS form 100 booklets) are readily available in the hospital |
|  | Who was to make the changes? | The QI team |
|  | Where were the changes effected? | All wards |
|  | For how long were the changes tested? | Two weeks |
| **Do** | After the above plan, we “do” the changes | All nurses |
| **Study** | What did we learn from the changes? | There was an improvement in the assessment of filling HMIS form 100 as shown in DHIS2 |
| **Act** | After learning, from the changes, we acted | The changes were adopted |

### 2.11 Sustaining changes

The team sustained routine reviews of the deaths, mentorship of student nurses on filling out HMIS form 100 following death on the ward, and left reminders at the nurses’ workstation. We proposed to introduce the changes by making them the new standard for service delivery during the project period and beyond and by sharing results widely to influence practice nationally through advocacy as an additional way to sustain changes.

### 2.12 Intervention monitoring and evaluation

We developed two process indicators and one outcome indicator from the sustained changes and prioritized the problem. The progress of the process indicators was tracked monthly while the outcome indicator was measured at the end of the six-month project period. It involved measuring the change in MCCoD and mortality reporting at MRRH. It involved collecting and analyzing quantitative data from DHIS2, hospital records, and death certificates. It also involved collecting and analyzing qualitative data from interviews with key stakeholders. The purpose of this phase was to evaluate the effectiveness and efficiency of the interventions in achieving the project objectives.

## 3.0 Results

### 3.1 Pre-intervention and baseline assessment

A team of 15 nurses and midwives was selected and trained in Continuous Quality Improvement (CQI), forming the CQI team. We appointed the Assistant Principal Nursing Officer to lead the project, coordinating meetings and serving as the liaison with the Principal Investigator. The composition of the QI team included ward in-charges, area managers from all departments, and at least one additional nursing staff member from each department.

At baseline, we found that a total of 141 deaths were reported in September 2023 but only 13 had been notified giving a notification rate of 9%.

### 3.2 Gaps identified during the baseline analysis

Gaps identified during the baseline analysis revealed three major challenges: recurrent stockouts of the Notification form, inadequate knowledge of how to use the Notification and certification forms, and failure to fill the HMIS form 100 correctly as soon as the death occurs.

### 3.3 Problem Prioritization results

The prioritization results indicate that the most pressing issue is the failure to fill the HMIS form 100 correctly as soon as a death occurs, scoring 18 points. This is followed by inadequate knowledge of how to use the notification and certification forms, which received a score of 17. Lastly, recurrent stockouts of the notification forms (HMIS 100) were identified as a significant concern, with a total score of 15 as shown in Table 2.

**Table 2.** Problem prioritization matrix for improving quality of Mortality notification at Mbale Regional Referral Hospital, 2022-2023.

| Problem | Important to the Next of keen<br>(1-5) | Affordable in terms of time and resources<br>(1-5) | Easy to measure<br>(1-5) | Under the control of team members<br>(1-5) | Total score |
| --- | --- | --- | --- | --- | --- |
| Recurrent stockouts of the Notification forms (HMIS 100) | 3 | 3 | 5 | 4 | 15 |
| Inadequate knowledge of how to use the Notification and certification forms | 4 | 4 | 4 | 5 | 17 |
| Failure to fill the HMIS form 100 correctly as soon as the death occurs. | 5 | 5 | 4 | 4 | 18 |

### 3.4 Root cause findings

The identified root causes included a lack of awareness among staff regarding the importance of timely form completion, which can lead to delays. Additionally, there was insufficient training on the procedures for completing the form, resulting in errors or omissions. Resource constraints, such as limited access to necessary time or personnel, further hindered prompt completion. Communication gaps among staff members also contributed to misunderstandings about responsibilities related to the form. Finally, broader systemic issues within the hospital exacerbated delays in filling out the HIMIS form as shown in Figure 1 below.

**Figure 1:**
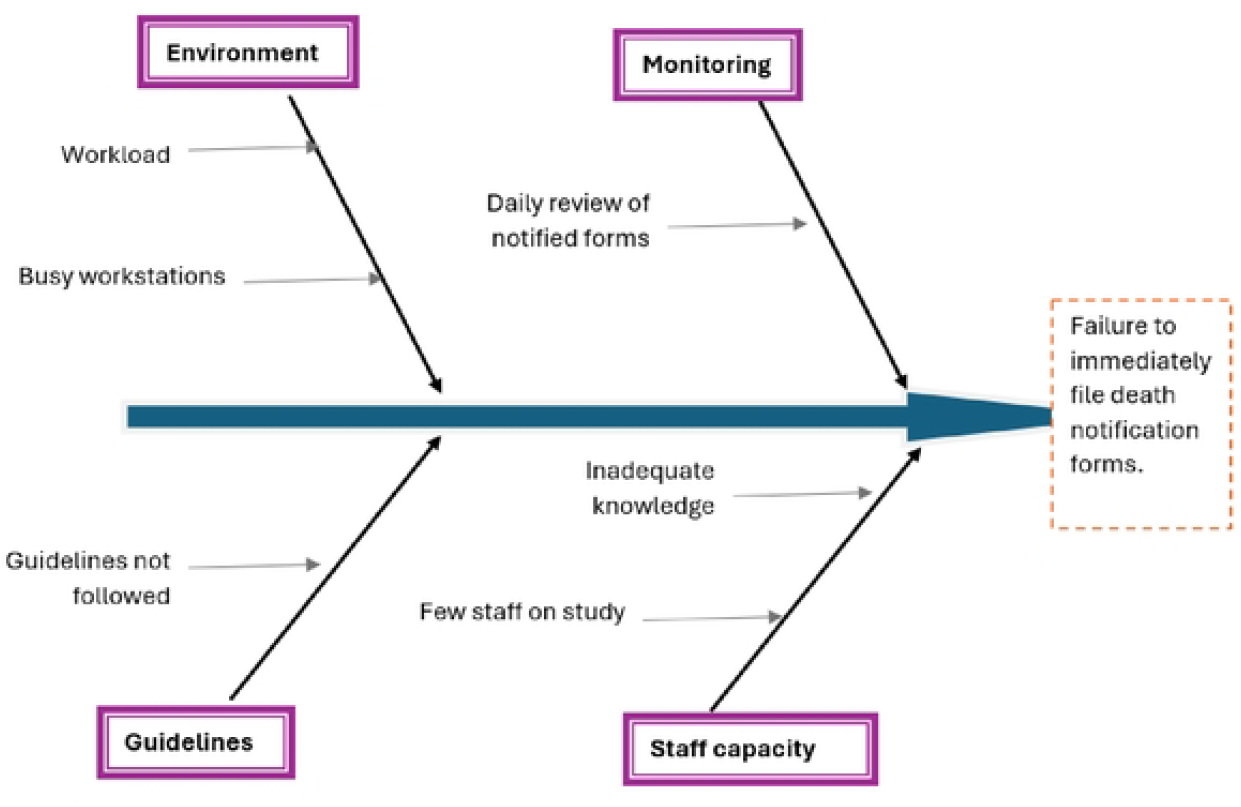
Root causes for failure of immediate filling of HIMIS form 100, Mbale RRH, 2023-2024.

### 3.5 Challenges in Completing HMIS Form 100

The group discussions revealed several key challenges that hindered the timely and accurate completion of the HMIS 100 death notification form, here we report direct quotes from the key stakeholders:

#### Lack of Knowledge and Training

Many nursing staff expressed a lack of understanding of how to properly fill out the various sections of the HMIS Form 100. They reported receiving limited training on the purpose and process of mortality reporting, leading to confusion and errors when attempting to complete the forms.

“I’ve never been trained to use the HMIS form 100. I’m unsure what information I should fill out and where. It’s all very confusing.”-Nurse, Mbale RRH

#### Competing Priorities and Workload

The nursing staff often cited their heavy workload and competing priorities as barriers to completing the HMIS Form 100 immediately after a patient’s death. The demands of providing direct patient care took precedence, leaving little time to focus on administrative tasks like mortality reporting.

“After a patient passes away, there are so many other urgent things I need to take care of. Filling out that form just gets pushed to the side because I’m busy with other patients.” - A Nurse, Mbale RRH

#### Lack of Supplies and Resources

Intermittent stockouts of the HMIS Form 100s were reported as a significant challenge, with nurses often unable to access the necessary forms when needed. This disrupted the mortality reporting process and contributed to underreporting.

“Sometimes we run out of the HMIS form 100, and then we can’t fill them out when a patient dies. We must wait until new forms arrive, and by then it’s too late.”-Nursing Supervisor, Mbale RRH

#### Importance of Mortality Reporting Undervalued

Several respondents indicated that the importance of accurate and timely mortality reporting was not well understood or prioritized by hospital management and staff. This lack of perceived value undermined the motivation to ensure proper completion of the HMIS form 100.

“I don’t think the hospital leadership sees the value in this mortality reporting. They don’t emphasize it or hold us accountable for doing it properly.”-A Medical Records Officer, Mbale RRH

In summary, the qualitative findings highlighted the multifaceted challenges faced by hospital staff in completing the HMIS 100 death notification form, including knowledge gaps, workload pressures, resource constraints, and a lack of perceived importance of mortality reporting.

### 3.6 Monitoring Results

The monitoring results for increasing the number of HMIS form 100 filled immediately after death at Mbale Regional Referral Hospital from October 2023 to April 2024 show that 67% of the target for mentorship sessions was achieved, with 4 out of the planned 6 sessions conducted. In terms of review meetings, only 58% of the target was met, with 7 out of 12 planned meetings held. Regarding the output indicator, 32 nurses were mentored on filling the HMIS form 100, representing 57% of the target of 15 nurses per session. Finally, the outcome indicator revealed that 65% of deaths were notified immediately using HMIS 100, falling short of the 80% target. As shown in the table 3 below.

**Table 3.** Indicator monitoring matrix for the progress towards increasing the number of HMIS form 100 filled immediately after death at Mbale RRH, between October 2023 to April 2024.

| Input indicators | Target | Data source | Achieved |
| --- | --- | --- | --- |
| 1. Number of mentorship sessions conducted among the nurses on filling HMIS form 100 | One session every month (6 sessions) | Attendance sheets and the session minutes | 4 (67%) |
| 2. Number of review meetings on filling of HMIS form 100 during bi-weekly QI meetings | Two review meetings per month for 6 months (12 meetings) | Attendance sheets and the session minutes | 7 (58%) |
| **Output indicators** |  |  |  |
| 1. Number of nurses mentored on filling HMIS form 100. | 15 nurses per mentorship | Attendance sheets | 32 (57%) |
| **Outcome indicator** |  |  |  |
| The proportion of deaths immediately notified using HMIS 100 between September 2023 to April 2024 | 80% of deaths notified immediately after death | Daily morning review meeting notes. | 65% |

Following the mentorships and documentation reviews, the proportion of deaths notified in DHIS2 significantly increased from 18% in September 2023 to 65% in April 2024 (p=0.009) as shown in Figure 2 below.

**Figure 2:**
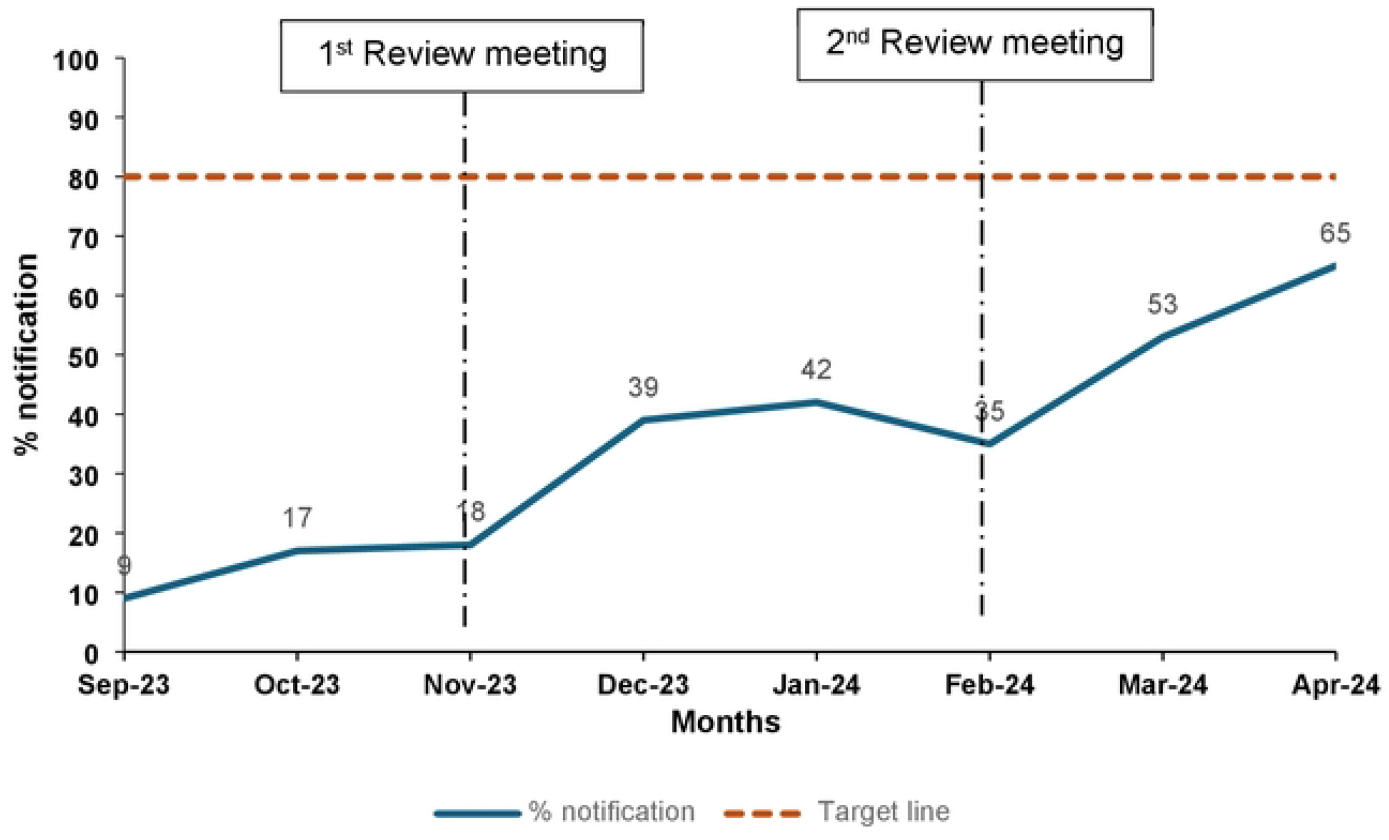
The trend of mortality notification from September 2023 to April 2024, DHIS2, Mbale Regional Referral Hospital.

## 4.0 Discussion

This quality improvement project aimed to increase the proportion of deaths occurring at Mbale Regional Referral Hospital that are notified through the DHIS2 system, from a baseline of 17% to a target of 80% over a 6-month period. The key interventions implemented included conducting staff mentorship sessions on properly completing the HMIS 100 death notification form, developing a standardized process for form completion, and ensuring the availability of necessary supplies and the results demonstrate a significant improvement in the proportion of deaths notified, increasing from 18% at baseline to 65% by the end of the project period.

The implementation of targeted interventions, such as the staff mentorship sessions and the development of a standardized process, helped to address some of these challenges. By equipping the nursing staff with the knowledge and tools to accurately complete the HMIS form 100, and by integrating mortality reporting into the hospital’s routine practices, the project was able to drive notable improvements in notification rates.

This progress, while still below the 80% target, marks a significant step forward in enhancing mortality reporting at the hospital following the staff training. A study done in US found that training staff on using a system in death notification positively impacts their ability to report on death [9]. However, this suggests that additional, more sustained efforts may be required to fully overcome the deeply entrenched barriers to effective mortality reporting. Continued investment in staff training [10], the consistent availability of necessary supplies, and the ongoing reinforcement of the importance of mortality reporting by hospital leadership will be crucial to maintaining and further enhancing the progress made. One study done in South Africa found that such continued efforts also minimize the number of errors that can be found in the notification reports [11].

The qualitative findings provided insights into the factors that have historically hindered effective mortality reporting at the hospital. Consistent with our findings, a study conducted in New York identified negative attitudes and a lack of knowledge among emergency department staff as major limitations to death notification [12]. Similarly, in our work in Mbale, we analyzed potential root causes, allowing the team to develop a multi-pronged systemic approach to address the issue and formulate potential solutions. Other studies have shown that such comprehensive approaches can lead to significant improvements [13, 14].

Furthermore, the lessons learned from this project—such as identifying knowledge gaps and addressing attitudes—could have broader implications for enhancing mortality reporting across Uganda’s healthcare system. The interventions implemented, including training sessions and integrating mortality reviews into routine nurses’ meetings, could serve as a model for adaptation and replication by other regional referral hospitals, ultimately contributing to the strengthening of the country’s civil registration and vital statistics (CRVS) system.

## 5.0 Study limitations

The quality improvement project was carried out over a relatively short 6-month period. While this timeframe allowed for the implementation and initial evaluation of the interventions, a longer study duration would have provided more time to fully embed the changes and assess their long-term sustainability. The short project period may have limited the ability to achieve the full 80% target for mortality notification.

The project was implemented solely at Mbale Regional Referral Hospital, which limits the generalizability of the findings to other healthcare facilities in Uganda. Factors specific to the context of Mbale RRH, such as resources, staffing, and organizational culture, may have influenced the success of the interventions.

Expanding the project to multiple regional referral hospitals could provide broader insights into the challenges and solutions for improving mortality reporting.

The qualitative data collected through stakeholder interviews relied on self-reported experiences and perceptions of the hospital staff. While this provided valuable insights into the barriers to mortality reporting, self-reported data can be subject to social desirability bias and may not fully capture the complexity of the issues.

Incorporating additional observational data or document reviews could have strengthened the analysis.

## 6.0 Conclusions

The quality improvement project at Mbale Regional Referral Hospital successfully increased mortality reporting through the HMIS 100 death notification form from a baseline of 18% to 65% over a 6-month period, highlighting the effectiveness of targeted interventions in addressing the multifaceted barriers to effective mortality reporting, including gaps in staff knowledge and training, competing priorities and heavy workloads, resource constraints, and the perceived lack of importance placed on this critical data collection process.

## Data Availability

The datasets upon which our findings are based belong to the Uganda Public Health Fellowship Program. For confidentiality reasons, the datasets are not publicly available. However, the data can be made available upon reasonable request from the corresponding author (Innocent Ssemanda) with permission from the Uganda Public Health Fellowship Program.

## List of abbreviations

MCCoD: Medical Notification and Certification of Cause of Death
CRVS: Civil Registration and Vital Statistics;
MoH –: Ministry of Health;
NSSF: National Social Security Fund;
DHIS2: District Health Information System version 2;
WHO: World Health Organization;
ICD: International Classification of Disease;
CQI: Continuous Quality Improvement;
PDSA: Plan-Do-Study-Act;
HMIS: Health Management Information System;
MRRH: Mbale Regional Referral Hospital;
QI: Quality Improvement;
CME: Continuous Medical Education

## Declarations

### Ethics approval and consent to participate

A non-research determination form was submitted to the US CDC for clearance before the commencement of the study. The Office of the Associate Director for Science, U.S. Centers for Disease Control and Prevention approved the study as a non-research project. This activity was reviewed by CDC and was conducted consistent with applicable federal law and CDC policy. §§See e.g., 45 C.F.R. part 46, 21 C.F.R. part 56; 42 U.S.C. §241(d); 5 U.S.C. §552a; 44 U.S.C. §3501 et seq. The study used data without capturing unique identifiers of the deceased with permission from the Ministry of Health and Mbale Regional Referral Hospital.

## Funding and disclaimer

This study was supported by the President’s Emergency Plan for AIDS Relief (PEPFAR) through the United States Centers for Disease Control and Prevention Cooperative Agreement number GH001353-01 through Makerere University School of Public Health to the Uganda Public Health Fellowship Program, Ministry of Health. The contents of this manuscript are solely the responsibility of the authors and do not necessarily represent the official views of the US Centers for Disease Control and Prevention and the Department of Health and Human Services, Makerere University School of Public Health, or the Uganda Ministry of Health.

## Authors’ contributions

IS: participated in the conception, investigation, design, analysis, and interpretation of the study and wrote the draft manuscript; ED and CN participated in the development of the concept and review of the results; JM supervised the project implementation and participated in the development of the concept and review of the results; BK and LB reviewed the report, reviewed the drafts of the manuscript for intellectual content and made multiple edits to the draft article; SO reviewed the report and ensured data accuracy: RM, LB and ARA reviewed the final manuscript to ensure intellectual content and scientific integrity. All authors read and approved the final bulletin.

## Acknowledgments

The authors would like to thank the Ministry of Health and the Mbale Regional Referral Hospital administration for their permission and support in accessing the hospital premises, staff, and data utilized in this project. We would also like to thank the staff at the Uganda National Institute of Public Health for their technical assistance.

